# Impaired speaking-induced suppression predicts degraded agency and hallucination severity in schizophrenia

**DOI:** 10.1101/2024.09.30.24314623

**Authors:** Songyuan Tan, Yingxin Jia, Miriam Mathew, Namasvi Jariwala, Alvincé Pongos, Kurtis Brent, Judith Ford, Daniel Mathalon, John Houde, Srikantan Nagarajan, Karuna Subramaniam

**Affiliations:** Department of Psychiatry, University of California, San Francisco, CA; Department of Otolaryngology, University of California, San Francisco, San Francisco, CA; Department of Radiology and Biomedical Imaging, University of California, San Francisco, San Francisco, CA; Department of Psychology, Palo Alto University, CA; Department of Psychiatry, UCSF and Veterans Affairs San Francisco Healthcare System, San Francisco, CA

**Author notes:** Corresponding Author: Karuna Subramaniam.

## Abstract

**Background:** Agency is the awareness of being the originator of one’s own thoughts and actions. Patients with schizophrenia (SZ) show deficits in agency that contribute to distortions in reality-monitoring (distinguishing self-generated from externally-produced information) and result in psychotic symptoms. Agency is also critical for speech-monitoring (monitoring what we hear ourselves say while speaking). For example, disruptions in agency that manifest as hallucinations are thought to result from the misattribution of the source of patients’ inner thoughts/speech as external voices.

**Methods:** We used magnetoencephalography (MEG) to assay assess agency during reality-monitoring (RM) and speech-monitoring (SM) tasks. In healthy controls (HC) during SM, the auditory cortical (A1) response is smaller while speaking (speak condition) compared to listening to the same speech (listen condition). This is known as speaking-induced suppression (SIS) M100 response which is measured using MEG 100ms after speech onset.

**Results:** During RM, SZ (N=30) showed impairments in both self-agency (identification of self-generated information) and external-agency (identification of externally-produced information), compared to HC (N=30). During SM, SZ failed to enhance M100 A1 responses during the listen condition and suppress M100 A1 responses while speaking, revealing impaired SIS. Weakened SIS predicted worsening hallucination severity.

**Conclusions:** SZ showed degraded neural M100 responses in A1 during the listen condition which drove impaired suppression of M100 SIS during highly-predictable self-generated speech. Impaired SIS induced noisier auditory sensory predictions, making it more likely for SZ to misattribute the source of inner thoughts/speech as externally-derived, giving rise to disruptions in agency during RM and more severe hallucinations.

## Introduction

Schizophrenia is a serious neuropsychiatric disorder affecting ∼1% of the population. Patients with schizophrenia (SZ) show cardinal deficits in agency - the awareness of being the cause of one’s own thoughts and actions (Ford and Mathalon, 2012; Subramaniam, 2021; Synofzik et al., 2010). Impairments in agency contribute to distortions in reality-monitoring (i.e., impairments in distinguishing self-generated from externally-produced events) and result in psychotic symptoms of hallucinations (Ford and Mathalon, 2012; Heinks-Maldonado et al., 2007; Subramaniam, 2021; Subramaniam et al., 2012; Subramaniam and Vinogradov, 2013; Synofzik et al., 2010). Agency is also critical for speech-monitoring (monitoring what we hear ourselves say while speaking). For instance, disruptions in agency that manifest as hallucinations occur when patients misattribute the source of their inner thoughts/speech as external voices (Ford and Mathalon, 2012; Ford et al., 2001a, b; Heinks-Maldonado et al., 2007; Subramaniam, 2021). Antipsychotic medications are far from adequate with up to 40% of SZ who continue to remain symptomatic (Lowe et al., 2018). For example, although clozapine is considered the most efficacious antipsychotic medication in refractory patients, 40–70% of these patients achieve only partial response to it while suffering from the side effects of antipsychotic medication (Nathou et al., 2019). Thus, there is an urgent need to understand the neural impairments underlying agency deficits that drive psychotic experiences in SZ.

In the present study, we used magnetoencephalography (MEG) to assess agency during reality-monitoring (RM) and speech-monitoring (SM) tasks. Our prior studies reveal that healthy control participants (HC) showed increased activity in the medial prefrontal cortex (mPFC) during the successful encoding and retrieval of self-generated information on our RM task which correlated with accurate judgments of self-agency, indicating that mPFC represents one neural correlate of self-agency (Subramaniam, 2021; Subramaniam et al., 2019). In other words, the more HC activated mPFC during encoding of self-generated information, the more mPFC activity better accurate retrieval of this self-generated information. By contrast, while performing the RM task in our fMRI studies, SZ showed hypoactivation in mPFC and left posterior superior temporal gyrus/sulcus (L.pSTG/S) associated with impairments in both self-agency judgments (i.e., during identification of self-generated information) and external-agency judgments (i.e., identification of externally-produced information), compared to HC (Subramaniam, 2021; Subramaniam et al., 2012; Subramaniam et al., 2017). We now examine whether we can extend these fMRI findings using MEG in a different sample of HC and SZ, and for the first time, we examine whether there is a common underlying mechanism that underlies impairments in SM and RM tasks, that results in psychotic symptoms in SZ.

In HC normally during SM, the primary auditory cortical (A1) response in L. pSTG/S is smaller while speaking (speak condition) compared to listening to the same speech (listen condition). This is known as speaking-induced suppression (SIS) M100 response which can be measured using MEG about 100ms after speech onset (Chang et al., 2013; Ford and Mathalon, 2004, 2012; Whitford, 2019). In other words, in HC, self-generated (and thus highly predictable) sounds give rise to suppressed responses (SIS), thus allowing speakers to pay better attention to sounds in the external world(Chang et al., 2013; Ford and Mathalon, 2004, 2012; Whitford, 2019). SIS is therefore indicative of highly-predictable ‘self-generated forward models’ (also known as efference copies/corollary discharge neural mechanisms) that allow all animal species to discount sensations resulting from their own actions(Ford and Mathalon, 2012; Poulet and Hedwig, 2006; Salomon and Starr, 1963). In other words, SIS is the phenomena that in HC, self-generated speech is highly predictable, and therefore elicits a suppressed response in left A1, compared to external speech(Chang et al., 2013; Ford and Mathalon, 2012; Whitford, 2019), indicative of a biological basis for self-agency that is essential for normal interactions with outside reality(Ford and Mathalon, 2012; Poulet and Hedwig, 2006; Salomon and Starr, 1963). By contrast, prior studies have shown that SZ reveal impaired M100 A1 SIS responses (i.e., impaired M100 while speaking compared to listening to the same speech) (Ford and Mathalon, 2004; Heinks-Maldonado et al., 2007; Whitford, 2019). Here, we now examine the mechanisms that drive such a weakened M100 SIS response in SZ, and for the first time we examine how impairments in low-level M100 SIS responses during SM relate to higher-level agency judgments during the RM task, and result in psychotic symptoms of hallucinations in SZ.

We had three specific hypotheses: (1) During the RM task, SZ would show impairments in both self-agency judgments (i.e., impaired identification of self-generated information) and external-agency judgments (i.e., impaired identification of externally-produced information), compared to HC. (2) During the SM task, SZ would show SIS impairments, revealed by a weakened M100 response during listen condition compared to speak (i.e., impaired M100 listen response minus M100 response during the speak condition). (3) SIS impairments in SZ would predict agency impairments during RM and worsening hallucination severity. If these hypotheses are confirmed, the present study would provide the first evidence for impairments in agency during a high-level cognitive judgment-based RM task that are driven by M100 primary auditory sensory impairments during a low-level SM task that provides a unitary basis for the underlying mechanisms that result in psychotic symptoms of hallucinations in SZ.

## Methods

### Participants

The present MEG study constitutes the baseline MEG portion of an NIMH-funded R01 (R01MH122897) study in schizophrenia to Karuna Subramaniam. Participants were recruited through our clinicaltrials.gov site (NCT04807530) or from our prior research studies if they had provided consent to be contacted for future studies. In addition, SZ were recruited from community mental health centers and outpatient clinics, and HC were recruited via advertisement. SZ (N=30) and HC (N=30) were matched at a group level on age and gender, and provided informed consent for this IRB-approved protocol. Participants then completed this MEG study and all clinical assessments at the University of California San Francisco (UCSF) (see Table 1). Inclusion criteria for SZ were Axis I schizophrenia diagnosis, assessed with the DSM-V [SCID] or, for HC, no psychiatric disorder, no substance abuse, meets MRI criteria, age between 18 and 64 years, right-handed, and English as the first language. Two HC and 2 SZ were either ineligible/unavailable to complete the MEG portion of the study. Hallucination severity was assayed using the Scale for the Assessment of Positive Symptoms (SAPS) (Andreasen et al., 1995).

**Table 1.** Demographic and Clinical Profiles (mean, SD) of Healthy Control (HC) and Schizophrenia Participants (SZ)

|  | **HC**<br>**(N=30)** | **SZ**<br>**(N=30)** | **P-value** |
| --- | --- | --- | --- |
| **Age (years)** | 40 (15) | 38 (14) | .60 |
| **Gender** | 20M, 10F | 21M, 9F | .78 |
| **SAPS Hallucination Severity** | N/A | 2.6 (1.9) | N/A |
| **Chlorpromazine (CPZ)**<br>**Equivalents** | N/A | 254 (240) | N/A |
| **Illness Duration (years)** | N/A | 11.6 (10.2) | N/A |

### Reality Monitoring Task

Participants in the study performed RM and SM tasks in the MEG scanner. As described previously (Subramaniam, 2021; Subramaniam et al., 2018a; Subramaniam et al., 2020; Subramaniam et al., 2018b; Subramaniam et al., 2012), the RM task consisted of an encoding phase and a memory retrieval phase (Figures 1A-B). All participants completed eight runs, with 20 trials per run, totaling 160 trials for the whole task. During the encoding phase, participants were presented with semantically-constrained sentences with “noun-verb-noun” format. On half of the sentences, the final word was either left blank for participants to generate themselves (e.g., *The <u>stove</u> provided the* _____), or was externally-generated by the experimenter (e.g., *The <u>sailor</u> sailed the <u>sea</u>*) (Fig. 2A). For each sentence, participants were told to pay attention to the underlined nouns for a subsequent memory test and to vocalize only the final word of each sentence. After the encoding phase, participants then completed the memory retrieval phase where they were randomly presented with the underlined noun pairs from the sentences (e.g., *stove-heat*), and were asked to identify by pressing a button on the button box whether the second word was previously self-generated or externally-generated. Self-agency judgments were computed as the number of correctly-identified self-generated trials, and external-agency judgments were computed as the number of correctly-identified externally-derived trials.

**Figure 1.**
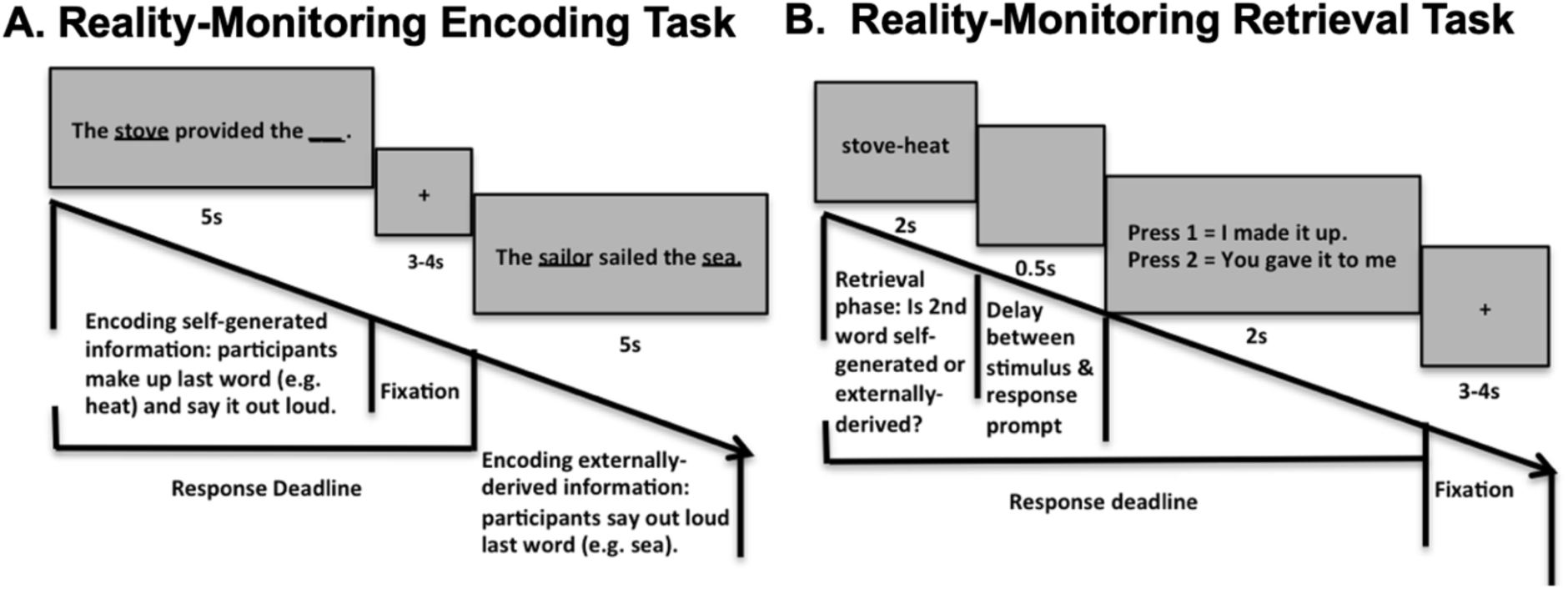
(A-B) Reality-Monitoring Task. **A.** Encoding: Participants are given semantically constrained sentences where on half of the sentences, the final word is left blank for SZ to generate themselves (e.g. *The <u>stove</u> provided the __*), and on the remaining half, the final word is externally-derived as it is provided by the experimenter (e.g.*The <u>sailor</u> sailed the <u>sea</u>*). Participants vocalize the final word of each sentence. **B.** Retrieval: Participants are randomly presented with the noun word pairs from the sentences, and need to identify with a button-press whether the 2^nd^ word was previously self-generated (e.g., *stove-heat*) (i.e., make self-agency judgments), or externally-derived (e.g., *sailor-sea*) (i.e., make external-agency judgments). We assess neural activity underlying agency by comparing MEG activity during encoding & retrieval of self-generated information with externally-derived information.

**Figure 2.**
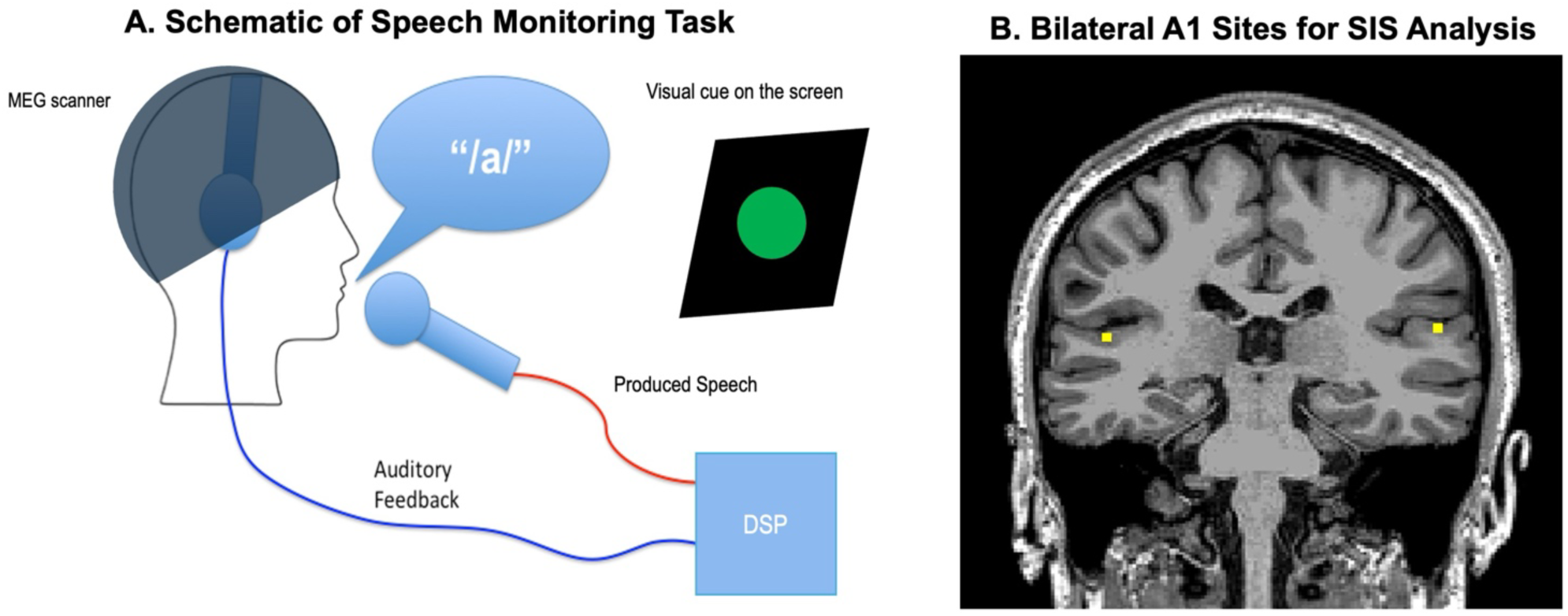
**A**. During the MEG speak session, participants vocalize the vowel /ɑ/ when they see a green dot on the screen, while listening to real-time auditory feedback in headphones via the digital signal processor (DSP). During the MEG listen session, when the green dot appears on the screen, participants listen to their recorded phonation of the vowel /ɑ/ from the speak session. **B.** We use MEG to record the M100 response based on MRI coordinates of primary auditory cortex (A1) in each subject (see yellow dots in D) (LH MNI: x,y,z=-54, -27, 12; RH MNI: x,y,z=54, -27,12). M100 responses are smaller during self-generated speech (i.e., saying the vowel /ɑ/), compared to hearing this same speech in subsequent playback. This is known as speaking induced suppression (SIS) response.

### Speech Monitoring Task

During the MEG SM task, participants wore a MEG-compatible microphone (AKG Pro Audio C520 Professional Head-Worn Condenser Microphone, Austria) and a pair of headphones. The microphone was connected to an amplifier, which, in turn, was connected to a Dell computer sound card (M-Audio Delta 44 4x4 analog I/O, M-Audio, Cumberland, RI). The amplified audio signal was transmitted back to participants through headphones. Before the experiment, participants confirmed that they could clearly hear the audio signal.

Participants completed two sessions, speak and listen, where each session consisted of 9 runs constituting 15 trials per run, totaling 135 trials. Each trial began with a visual cue (i.e., a green dot) that appeared on the screen. For the Speak session, participants were instructed to vocalize the vowel /ɑ/ when they saw the green dot. They continued the phonation for 2.5 seconds until the dot disappeared while listening to real-time auditory feedback from the headphones. In the listen session, participants passively listened to their own recorded phonation playback from the Speak session when they saw the green dot on the screen. The inter-trial interval was 2.5s during which time participants viewed a blank screen.

### MRI Acquisition

Structural T1-weighted MRI images were obtained for each participant using a 3-Tesla Siemens MRI scanner at the UCSF Neuroscience Imaging Center for MEG source space reconstruction. For each subject, high-resolution MRI was acquired using an MPRAGE sequence (160 1-mm slices, field of view = 256 mm, repetition time = 2300 ms, echo time = 2.98 ms). For each subject, the outline of the brain on the structural scans was extracted, and the segmented brain was treated as a volume conductor model for the source reconstruction described below.

### MEG Data Acquisition

Magnetic fields were recorded using a whole-head 275-axial-gradiometer MEG system (MEG International Services Ltd., Coquitlam, BC, Canada) in a shielded room. The sampling rate was 1200 Hz with a 0.001-300 Hz bandpass filter. Three fiducial coils (nasion, left, and right preauricular points) were placed to localize the head position relative to the sensors. Three fiducial coils (nasion and left and right preauricular) were placed to localize the position of the head relative to the sensor array for co-registration of the MEG data with each individual’s structural anatomical MRI. Head localization was performed at the beginning and end of each task block to register head position and to measure head movement during the task. Third gradient noise correction filters were applied to the data and corrected for a direct current offset based on the whole trial. Noisy sensors and trials with artifacts (i.e., due to head movement, eye blinks, and saccades or sensor noise) were defined as magnetic flux exceeding 2.5 pT. epochs were rejected from further analysis if they contained artifacts.

### MEG Analyses

An analysis interval of 600ms was extracted for each trial, which spanned from -300ms to 300ms relative to the phonation onset. For each participant, the M100 response was averaged across all trials for each condition and each hemisphere. Co-registration of the MEG data with each individual’s structural anatomical MRI was performed based on the nasion, left, and right preauricular fiducial coil positions utilizing the Neurodynamic Utility Toolbox for MEG (NUTMEG; available at http://nutmeg.berkeley.edu). Bayesian covariance beamforming was applied to the average M100 response across all trials for left and right sensor arrays, focused on the left (MNI x,y,z = [-54.3, -26.5, 11.6]) and right A1 cortices (MNI x,y,z = [54.4, -26.7, 11.7]) in each participant (Kort et al., 2014). Source timeseries was converted into absolute Bayesian Covariance Beamformer (BCB) activity using Champagne (Bhutada et al., 2022; Cai et al., 2021) at each time-point from -100ms to 300ms, for a total duration of 400ms in left and right A1 using the MNI subject-specific coordinates for each person. The extracted time-course for each condition (speak and listen) in each hemisphere were normalized as z-scores for each participant. M100 responses were computed 100ms after speech onset (i.e., 50ms before and after the 100ms time-point after speech onset) during the listen condition and the corresponding speaking condition (Kim et al., 2023; Kort et al., 2014). To compute SIS, M100 responses were subtracted for listen compared to speak (i.e., M100 listen minus M100 speak response) at each time-point during this 100ms time-window for each participant in each hemisphere.

### Statistical analyses

One-way ANOVAs were implemented to examine group differences between HC and SZ, in M100 responses during SM as well as in self-agency judgments (i.e., self-generated identification accuracy) and external-agency judgments (i.e., externally-generated identification accuracy). Pearson’s two-tailed correlation tests were used to measure the strength of the linear relationship between M100 responses with agency judgments and hallucination severity.

## Results

Between-group one-way ANOVAs indicated that on the RM task, SZ revealed significant impairments on both self-agency judgments (F=4.99, p=.03) and external-agency judgments (F=7.38, p=.01), compared to HC. During the SM task in the listen session when participants were passively listening to their recorded phonation from the speak session, between-group one-way ANOVAs revealed that M00 A1 activation in left pSTG/S in the SZ group was significantly lower than HC around 100ms after playback onset (F=4.23, p=.045) (Figure 4A). Specifically, SZ showed reduced left M100 responses in the time-range from 90 to 110ms. This reduced M100 response in the listen condition drove a reduced M100 SIS (i.e., listen minus speak) response (F=4.39, p=.041) within the same time-range of 90 to 110ms (Figure 5A). We did not find any between-group differences in M100 response in the right hemisphere or during the Speak condition (all p’s>.80).

**Figure 3.**
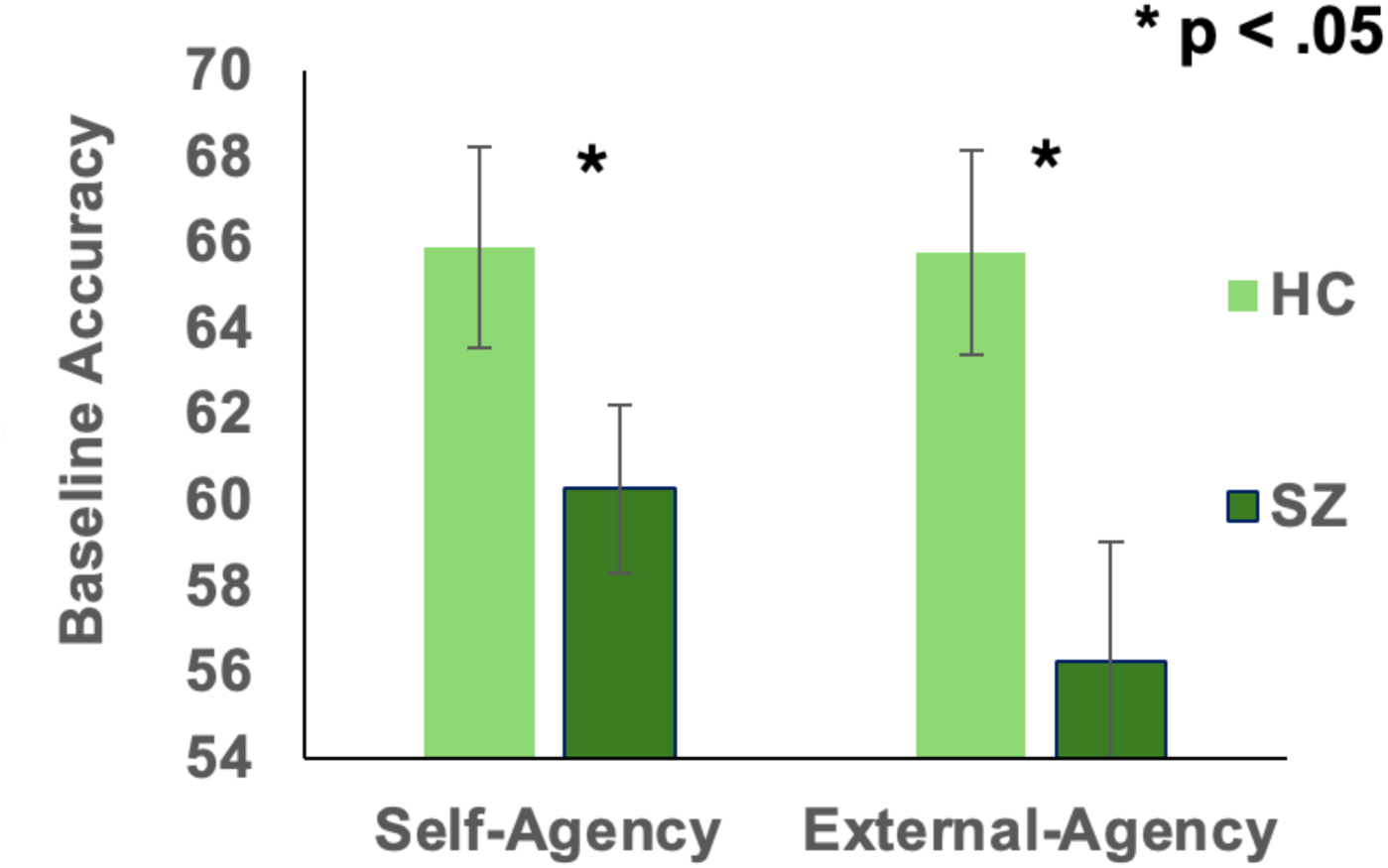
SZ showed impairments in agency during performance on the reality-monitoring task compared to HC.

**Figure 4.**
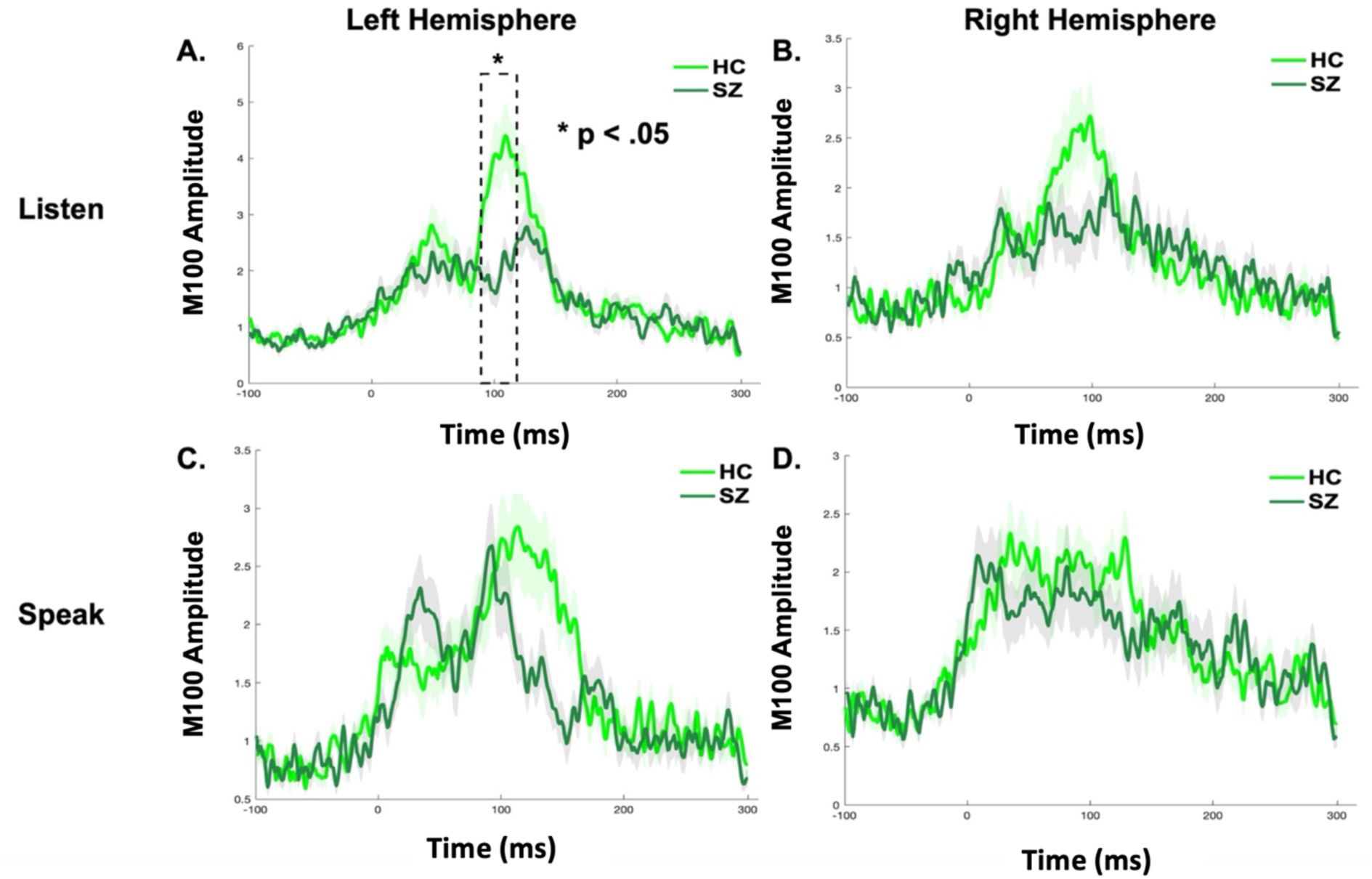
**(A-D).** Compared to HC, SZ failed to enhance neural M100 responses in left primary auditory cortex (A1) only during the MEG listen condition.

**Figure 5.**
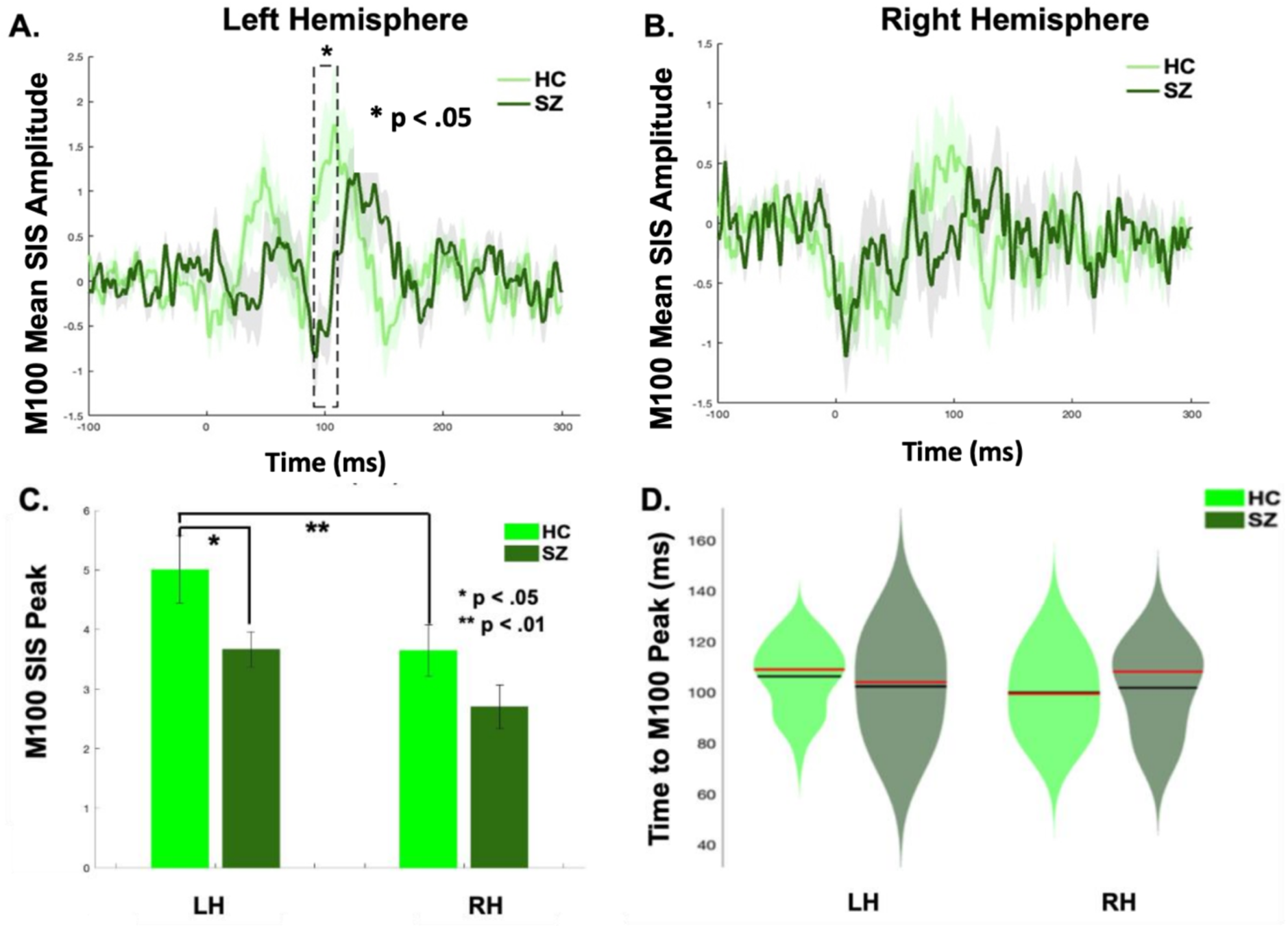
**A-B.** SIS mean amplitudes (z-scores) in left and right hemispheres were calculated by subtracting the M100 auditory evoked response during the speak condition at each time point from the listen amplitude (i.e., SIS Amplitude = listen – speak). The black dashed box indicates the time window that showed a significant SIS difference about 100ms after speech onset (i.e., 90-110s) only in the left hemisphere between HC and SZ groups (p < .05). 0ms indicates voice feedback onset in speak and listen sessions. **C**. Bar charts illustrate a laterality effect in which HC reveal a significantly larger M100 peak SIS amplitude (i.e., peak listen minus speak z-scores) in the left hemisphere compared to the right hemisphere (**p<.01). This laterality effect is absent in SZ. HC also show a significantly great M100 SIS response in the left hemisphere, compared to SZ (*p< .05). **D**. The violin chart shows that there is no between-group difference or laterality effects in time to reach the M100 peak (median=red line; mean=black line).

We also found that the HC group revealed a laterality effect with significantly greater M100 A1 activation in left pSTG/S during the listen condition in the left hemisphere, compared to the right hemisphere (t=2.92, p=.007). Greater laterality M100 A1 responses in HC during the listen condition in the left hemisphere drove a greater M100 SIS (i.e., listen minus speak) laterality response in the left pSTG/S, compared to the right pSTG/S (t=2.81, p=.009) (Figure 5C). This laterality effect was not observed in the speak condition in HC or in SZ in any condition (i.e., listen, speak or SIS conditions) (all p’s>.05). We did not find any laterality effects in the time to reach peak M100 A1 responses in either group, nor did we find any between-group differences in the time to reach peak M100 response (all p’s>.05) (Figure 5D).

Interestingly, we found that SZ who showed greater M100 responses in left pSTG/S during the listen condition while performing the SM task, also revealed greater external-agency judgments during the RM task (r=.39, p=.04). This significant relation was also sustained for SZ who showed greater M100 SIS responses (i.e., listen minus speak) in left pSTG/S, which also predicted greater external-agency judgments during the RM task (r=.38, p<.05) (Figure 6). Finally, we found that SZ who showed greater M100 A1 responses in the left pSTG/S during the listen condition and during SIS, revealed lower hallucination severity on the SAPS scale (r=-.40, p=.03 and r=-.38, p<.05, respectively) (Figure 7).

**Figure 6.**
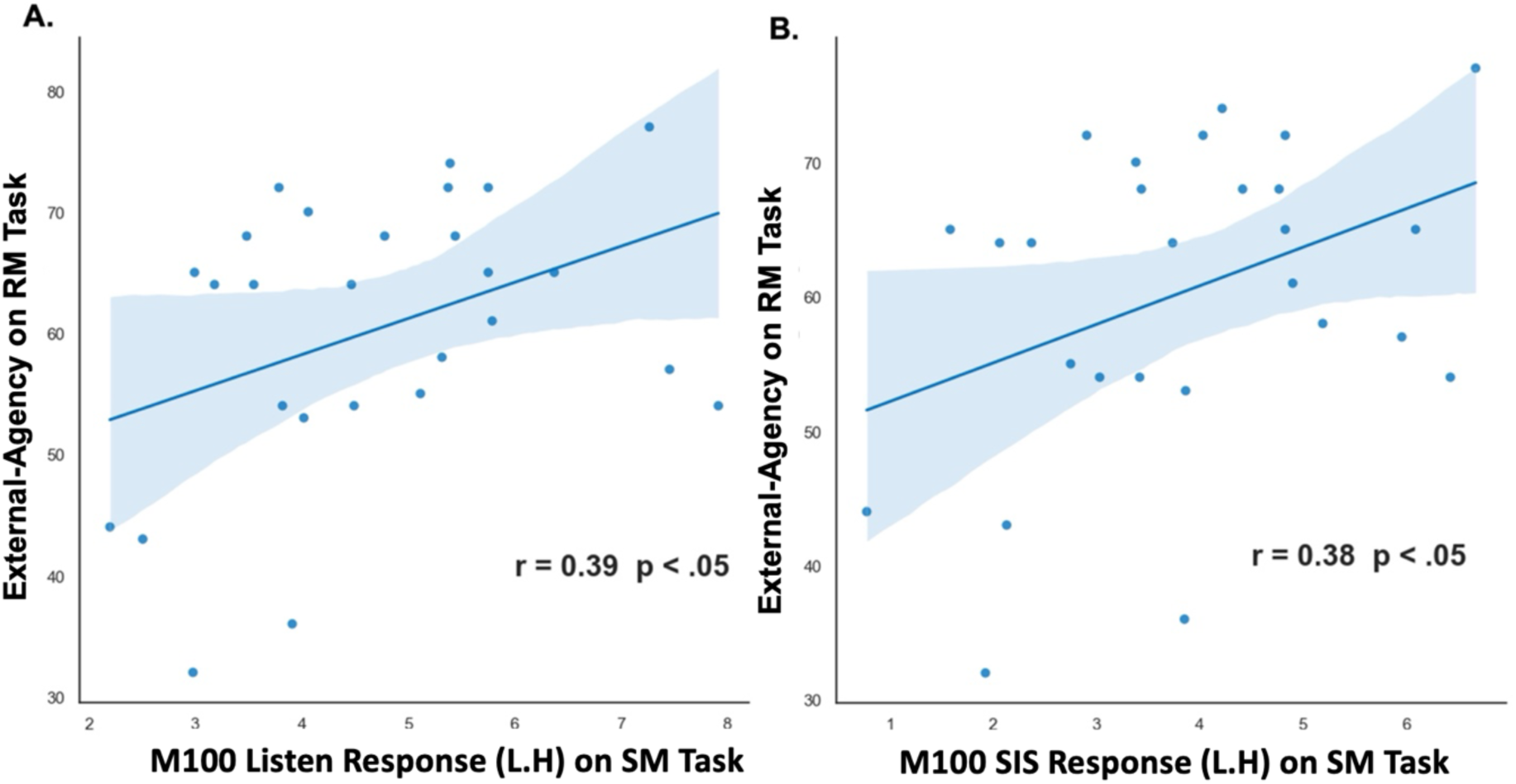
The scatter plots illustrate significant positive correlations between **(A)** MEG M100 listen and **(B)** M100 SIS (i.e., listen minus speak) responses in the left hemisphere during the SM task with external-agency during the RM task. The shaded area indicates 95% confidence intervals.

**Figure 7.**
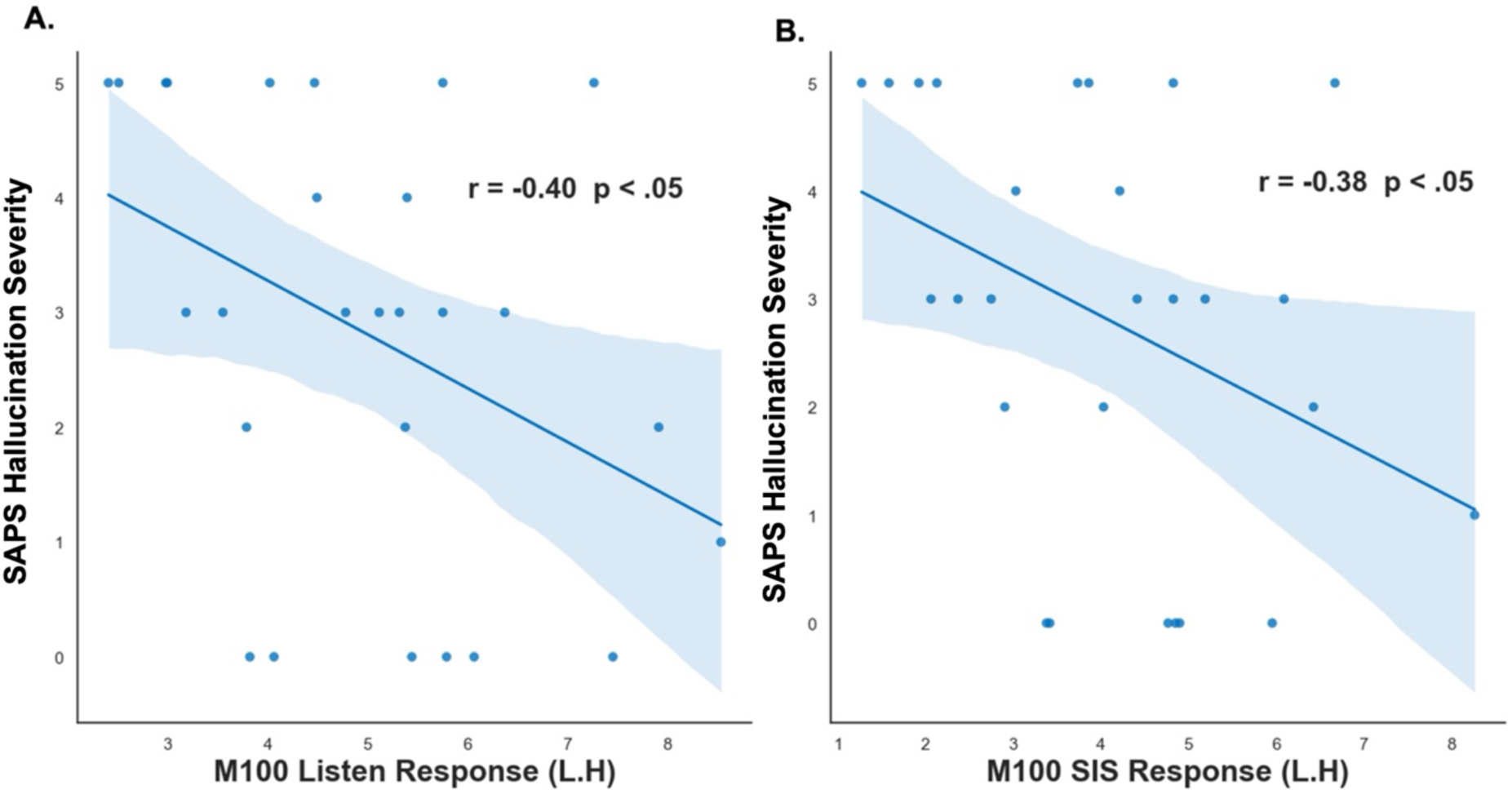
The scatter plots illustrate significant negative correlations between **(A)** MEG M100 listen and **(B)** M100 SIS (i.e., listen minus speak) responses in the left hemisphere during the SM task with hallucination severity on the SAPS scale. The shaded area indicates 95% confidence intervals.

## Discussion

In this study, we found that SZ had: (1) Impairments at both self-agency (i.e., identifying themselves as the originator of self-generated information) and external-agency (i.e., identifying externally-derived information) on the RM task, (2) Degraded M100 A1 responses in left pSTG/S during the listen condition, that contributed to impaired M100 A1 SIS responses, (3) SZ who had greater M100 responses in left pSTG/S during the listen condition and during SIS, also showed greater external-agency judgments, and finally (4) SZ who had greater M100 responses in left pSTG/S during the listen condition and during SIS also had lower hallucination severity. Together, these convergent findings across distinct tasks, reveal that the failure to enhance auditory sensory responses to information in the external environment on a low-level SM task suggests that SZ have noisier auditory sensory predictions that impaired their ability to identify externally-produced information on a high-level RM task, making them more likely to misattribute the source of self-generated information (i.e., inner thoughts/speech) as being externally-produced, giving rise to disruptions in agency judgments and hallucinations (Ford et al., 2007a; Ford and Mathalon, 2004, 2012; Ford et al., 2001a, b; Ford et al., 2007b; Heinks-Maldonado et al., 2007). This is the first proof-of-concept study to delineate the neural underpinnings of hallucination severity in SZ by relating auditory sensory impairments in SZ on a low-level SM task that predicted agency impairments on a completely different high-level RM task, which resulted in worsening psychotic hallucination severity.

Our findings in HC are consistent with prior research revealing that HC showed stronger laterality M100 responses in the left hemisphere during the listen condition and during SIS, concordant with left hemisphere dominance for language processing (Binder et al., 2000; Curio et al., 2000; Houde et al., 2002; Kort et al., 2014; Zatorre et al., 2002; Zatorre et al., 1992). This stronger laterality M100 response in the left hemisphere was not observed in the SZ group. We did not find any between-group differences in the time to reach peak M100 responses, but only in M100 amplitude responses only during the listen condition, but not in the speak condition, that were greater in HC compared to SZ, which were specific to the left hemisphere. Normally in HC, M100 responses while speaking gives rise to suppressed responses (SIS), thus allowing speakers to enhance M100 responses while listening to sounds in the external world(Chang et al., 2013; Ford and Mathalon, 2004, 2012; Whitford, 2019). SIS is therefore indicative of highly-predictable ‘self-generated forward models’ (i.e., efference copies/corollary discharge neural mechanisms) that allow all species across the animal kingdom to discount sensations resulting from their own actions(Ford and Mathalon, 2012; Poulet and Hedwig, 2006; Salomon and Starr, 1963). SIS thus provides a primordial basis for self-agency that is necessary for normal interactions with the outside world (Ford and Mathalon, 2012; Poulet and Hedwig, 2006; Salomon and Starr, 1963). In SZ, we found that this failure to enhance M100 auditory sensory responses to information in the external environment during the listen condition and suppress M100 responses while speaking suggests that SZ have noisier auditory sensory predictions, making them more likely to misattribute the source of inner thoughts/speech as being externally-produced, giving rise to disruptions in agency judgments on the high-level RM task and psychotic disruptions in reality-monitoring that resulted in hallucinations (Ford et al., 2007a; Ford and Mathalon, 2004, 2012; Ford et al., 2001a, b; Ford et al., 2007b; Heinks-Maldonado et al., 2007). These results were not due to differences in latency timing to reach peak M100 responses, but were only due to degraded M100 amplitude auditory sensory responses in the listen condition that were specific to the left hemisphere that is known to have dominance for language processing.

Our prior fMRI studies revealed that SZ showed hypoactivation in medial frontal and L.pSTG/S sites associated with impairments in both self-agency judgments and external-agency judgments on the RM task, compared to HC (Subramaniam, 2021; Subramaniam et al., 2012; Subramaniam et al., 2017). Here, we now replicate and extend these prior fMRI findings using MEG in a different sample of HC and SZ, and provide the first evidence for impairments in agency during a high-level RM task that is driven by degraded A1 M100 auditory sensory activity in L.pSTG/S during a low-level SM task, thus providing a unitary basis for the underlying mechanisms that result in psychotic symptoms of hallucinations in SZ. Prior studies using EEG have shown that medial frontal activity immediately prior to speech onset is strongly correlated with SIS response shown by N1 suppression (during speak minus listen conditions) about 100ms after speech onset (Ford and Mathalon, 2004; Ford et al., 2002). These data suggest that the brain generates efference copies (i.e., copy of the motor command) via medial frontal activity immediately prior to self-generated actions that represent highly-reliable predictions in ‘self-generated forward models’ that model the expected sensory outcome of self-generated actions during RM and SM tasks underlying agency(Chang et al., 2016; Franken et al., 2018; Khalighinejad et al., 2018; Subramaniam, 2021; Subramaniam et al., 2019). In HC, this medial frontal activity prior to vocalization is strongly correlated with subsequent suppression of auditory responses to the spoken sound in L.pSTG/S (Ford and Mathalon, 2004; Ford et al., 2002). By contrast, here, we found that SZ showed degraded neural M100 auditory sensory responses to information in the external environment in L.pSTG/S in the listen condition which drove impaired M100 SIS responses during highly-predictable self-generated speech (Ford and Mathalon, 2012; Ford et al., 2001a; Heinks-Maldonado et al., 2007). This resulted in noisier auditory sensory predictions, blurring the boundary between inner thoughts/speech and external sounds, giving rise to disruptions in agency judgments during RM and hallucinations in SZ.

In summary, we provide a novel perspective for investigating the underlying neural mechanisms underlying agency within two distinct speech monitoring and reality monitoring frameworks, and provide a unitary basis for external agency during a high-level RM task that results from the fundamental ability to sense and respond to external sounds during a low-level SM task (e.g. shown by enhanced M100 primary auditory sensory responses in L.pSTG/S). The present findings not only provide innovative functional biomarkers for understanding the underlying the neural basis of agency, but also provide the first step toward applying precision-medicine guided neuromodulation targets within medial frontal sites that we have previously found to underlie self-agency (Subramaniam, 2021; Subramaniam et al., 2019; Subramaniam et al., 2020; Subramaniam et al., 2012) and L.pSTG/S site that we show here mediates external-agency. Specifically, the present findings suggest that SZ will benefit from directly targeting and enhancing neural activity in L.pSTG/S that will augment auditory sensory responses to information in the external environment that will drive improved SIS while speaking, thus making it easier for SZ to distinguish the source of inner thoughts/speech from external speech and will potentiate reduced hallucinations. In conclusion, the present research creates a path towards developing new personalized neuromodulation treatment interventions that target medial frontal and L.pSTG/S sites to improve self-agency and external-agency, respectively, that will be particularly useful for patients with psychosis disorders who exhibit severe impairments in both self-agency and external-agency for maximally reducing hallucination severity.

## Data Availability

The data will be published in the NIMH Data Archive (NDA), as part of the NIMH Data Submission Agreement(DSA).

https://nda.nih.gov/

## Acknowledgments

We thank all the participants for completing our studies. This research is supported by NIMH R01 grants (R01MH122897 and R01MH122897-05S1) to Karuna Subramaniam.

